# Early Cancer Detection in Asymptomatic Subjects through Measurement of Crosslinked cf-Nucleosomes in Plasma

**DOI:** 10.1101/2025.03.14.25323908

**Authors:** Dorian Pamart, Marie Piecyk, Lea Payen, Guillaume Rommelaere, Brieuc Cuvelier, Adrien Govaerts, Marielle Herzog, Jacob Micallef

**Affiliations:** R&D Dept, Belgian Volition SRL, 22 Rue Phocas Lejeune, Parc Scientifique Créalys, 5032 Isnes, Belgium; Center for Innovation in Cancerology of Lyon (CICLY) EA 3738, Hospices Civils de Lyon, Faculty of Medicine and Maieutic Lyon Sud, Claude Bernard University Lyon I, 69921 Oullins, France; Department of Biochemistry and Molecular Biology, Lyon-Sud Hospital, Hospices Civils de Lyon, Pierre-Bénite, France; Volition Diagnostics UK Limited, 93-95 Gloucester Place, London, W1U 6JQ, UK

**Author notes:** **Corresponding Author:** Jake Micallef, Volition Diagnostics UK Limited, 93-95 Gloucester Place, London, W1U 6JQ, UK.

**Keywords:** nucleosome, chemiluminescent immunoassay, crosslinking, early cancer detection, solid cancers

## Abstract

**ABSTRACT:** *Purpose:* Plasma cell-free nucleosome (cf-nucleosome) measurements are used in veterinary, but not human medicine, to identify asymptomatic subjects at high risk of cancer. Uniquely among plasma sandwich immunoassay tests, cf-nucleosomes are nucleoproteins with a complex electrochemistry. Formaldehyde crosslinking is common in the investigation of cellular nucleoproteins by chromatin immunoprecipitation. cf-nucleosome sandwich immunoassay similarly involves isolation from plasma by chromatin immunoprecipitation. We investigated immunoassay of crosslinked plasma cf-nucleosomes in human cancer samples.

*Experimental Design:* Plasma cf-nucleosomes were covalently crosslinked by blood collection in tubes containing a formaldehyde-releasing agent. Plasma samples were collected from asymptomatic healthy volunteers, treatment-naïve cancer patients diagnosed with one of 21 different cancer types, and hospitalized patients with non-cancer conditions. Samples were assayed for a variety of nucleosome species at two independent sites and using immunoassays available from different commercial manufacturers. The assay cutoff was set at the highest cf-nucleosome level measured for any asymptomatic control subject to obtain results for cancer patients at 100% observed specificity.

*Results:* An elevated level of crosslinked plasma cf-nucleosomes was observed in 112 of 229 cancer patients tested (49% sensitivity), including 11 of 32 Stage I solid cancers (34% Stage I sensitivity). Elevated levels also occurred in hospitalized patients with non-cancer pathologies, but no false-positive results occurred among 150 asymptomatic control subjects.

*Conclusions:* Immunoassay of crosslinked plasma cf-nucleosomes achieved clinically meaningful sensitivity for cancer at 100% observed specificity in asymptomatic subjects, and shows promise for the identification of asymptomatic individuals at high risk of an undetected cancer.

**Significance:** Early detection of cancers by screening asymptomatic subjects through minimally invasive blood tests is a goal in oncology. We show that crosslinking of circulating cf-nucleosomes in human whole blood prior to immunoassay achieved clinically meaningful sensitivity for cancer (49% overall, 34% in Stage I disease) across multiple tumor types at 100% observed specificity among the asymptomatic controls tested, suggesting this technique may have broad utility for improving early cancer detection.

## Introduction

Cancer mortality varies greatly depending on whether the disease is detected at an early, localized stage when effective treatment options are more likely to be available, or at a later stage when the disease may have spread and treatment is more difficult. As cancer often becomes symptomatic only at later disease stages, patient outcomes are significantly improved by early cancer detection through screening of asymptomatic subjects (1). Current cancer screening methods, such as Fecal Immunochemical Testing (FIT) for colorectal cancer (CRC), low-dose computed tomography (LDCT) scanning for lung cancer, and mammography for breast cancer, are not diagnostic for cancer, but are minimally invasive, scalable, cost effective and useful for the identification of a subpopulation of asymptomatic subjects who are at elevated risk for cancer.

Cell-free nucleosomes (cf-nucleosomes) are the primary circulatory form of cell-free DNA (cfDNA) in the blood. Circulating cf-nucleosome levels are low in healthy subjects but may be elevated in patients diagnosed with cancer (particularly hematopoietic cancer), as well as a variety of other infectious or inflammatory conditions. Plasma cf-nucleosome levels measured by sandwich immunoassay are used in veterinary medicine for cancer screening in dogs. These tests have high (97%) specificity for cancer in asymptomatic dogs and are able to identify approximately half of all canine cancers (2, 3). Hematopoietic cancers represent almost 30% of canine cancers. cf-nucleosome levels are elevated in 82% of canine hemangiosarcoma cases, including 67% of dogs with Stage I disease, and 74% of canine lymphoma cases, including 67% of dogs with Stage I disease.

Sandwich immunoassays for circulating cf-nucleosomes are also reported to be promising in the management of human cancers (4). However, neither cfDNA nor cf-nucleosome levels have been used for early cancer detection in humans due to the low levels reported in early-stage solid cancers and the non-specific nature of nucleosomes as a biomarker (5, 6).

cf-nucleosomes are the only plasma nucleoprotein measured by sandwich immunoassay and their solution electrochemistry is unlike that of typical plasma protein analytes. Most plasma proteins are negatively charged at physiological pH, with isoelectric points in the range of pH 5–6. Proteins with a more acidic isoelectric point have a stronger net negative charge at physiological pH. For example, albumin is a strongly negatively charged plasma protein with an isoelectric point of pH 5 and a net negative charge of -17, which helps impede albumin binding to negatively charged cell-surface glycosaminoglycans (7). The net negative charge of an intact 147 base pair (bp) or 167 bp cf-nucleosome is -148 or -178, respectively. Neutralization of this charge by bivalent calcium and magnesium ions (Ca^2+^ and Mg^2+^) is central to their electrochemistry (8–13).

The nucleosome core particle (NCP) comprises a 147 bp DNA fragment bound to a protein core comprising two copies each of the histones H2A, H2B, H3, and H4. The 147 bp DNA component bears a negative charge of -294 (−2 per bp). The negative charge of the DNA is 20% neutralized by a +58 charge on the globular histone core. In intact nucleosomes, a further 33% of the negative charge is neutralized by a positive charge of +98 on the histone tails. The NCP is surrounded by a strong negative electrostatic field which is greatest at the edges of the nucleosome disc surrounding the coiled surface DNA and weaker around the histone core surfaces of the disc (8, 13).

In clipped nucleosomes, where one or more histone tails are removed, the net charge is increased by up to a further -98, making clipped nucleosomes less stable than intact nucleosomes (10, 14, 15). The conformation and stability of chromatin and cf-nucleosome structures are reported to be particularly influenced by electrochemical interactions with Ca^2+^ and Mg^2+^ ions, as well as by EDTA sequestration of those ions (9–12). Interestingly, other divalent cations such as zinc, iron, cobalt, or manganese ions (Zn^2+^, Fe^2+^, Co^2+^, and Mn^2+^) have little or no effect on the stability of histone-DNA interactions in clipped nucleosomes (10).

The stability of cf-nucleosomes is also dependent on the integrity of the associated DNA fragment. Introduction of single- or double-stranded DNA breaks or nicks using DNase or Nickase enzymes affects DNA superhelicity, destabilizes cf-nucleosomes and leads to a marked increase in histone eviction (16). The proportion of nicked cf-nucleosomes is reported to be consistent and low in the plasma of healthy subjects, but elevated in the plasma of cancer patients (17–19).

In addition to circulating in free solution, cf-nucleosomes are reported to circulate bound to the surface of blood cells through electrochemical interactions of positively charged histone tails with negatively charged glycosaminoglycans (20–23). Glycosaminoglycan binding is Mg^2+^ ion-dependent, and cell surface-bound cf-nucleosomes (csb-cf-nucleosomes) are reported to be eluted into free solution by EDTA sequestration of bivalent cations (10, 20).

Cellular chromatin immunoprecipitation (ChIP) studies commonly involve stabilization of nucleosomes and other nucleoproteins by crosslinking with formaldehyde prior to antibody binding to preserve nucleoprotein structures. Crosslinking renders nucleoproteins more stable to physical or chemical insult during biochemical procedures, leading to more robust results and better elucidation of weaker or less stable biological protein-DNA interactions than could otherwise be achieved (24–26). Although sandwich immunoassay methods for plasma cf-nucleosome measurement similarly involve nucleosome immunoprecipitation from plasma by antibody binding, immunoassay of crosslinked plasma cf-nucleosomes has not previously been described.

Some cfDNA blood collection tubes (BCT) contain formaldehyde-releasing agents to crosslink and stabilize the membranes of blood cells to prevent cell lysis and concomitant contamination of plasma with intracellular nuclear and mitochondrial DNA. The use of formaldehyde in these BCTs is intended to crosslink cell membranes, but formaldehyde is a non-specific reagent which crosslinks proteins and nucleoproteins more generally in these BCTs (27) and also crosslinks csb-cf-nucleosomes to cell membranes (22, 23). Studies of reaction kinetics show that formaldehyde crosslinking of nucleoproteins is rapid and complete within 5 seconds (24, 28).

Here we report results for the measurement of crosslinked cf-nucleosomes in the plasma of cancer patients and asymptomatic healthy volunteers by automated magnetic chemiluminescent immunoassay.

## Materials and Methods

### Native and crosslinked plasma samples

Native plasma samples were collected in EDTA plasma BCTs. Tubes were processed within 4 hours of venipuncture and separated plasma was stored frozen in cryotubes at - 80⁰C until assayed.

Covalent crosslinking of cf-nucleosomes in whole blood was performed by collection of samples in Streck Cell-Free DNA BCT tubes (Streck, La Vista, NE, USA) containing a formaldehyde-releasing agent. Samples were processed according to the manufacturer’s instructions. Separated Streck plasma was stored frozen in cryotubes at -80⁰C until assayed.

A power calculation using the Student’s t-test estimated the minimum sample size required to obtain results significant at the p<0.05 level as approximately 45 test patients and 45 control subjects (power = 80%, β = 0.2), assuming a difference between groups of 0.6 standard deviations.

Samples were collected from 598 subjects, including 389 treatment-naïve patients diagnosed with one of 21 different cancer types, 150 self-declared asymptomatic healthy volunteers, 49 hospitalized patients with elevated levels of the inflammatory marker C-reactive protein (CRP >5 mg/mL) and 10 non-hospitalized patients diagnosed with an inflammatory disorder (not in a flare). To monitor for potential center bias in sample collection, samples were obtained from multiple independent sources, including from commercial biobanks BioIVT (cancer, healthy volunteers, and non-hospitalized inflammatory diseases), Indivumed (cancer and healthy volunteers), INO Specimens Biobank (hospitalized patients with elevated levels of CRP), Etablissement Français du Sang (healthy volunteers), and from Centre Hospitalier Lyon Sud (cancer samples) collected in routine use.

Matched EDTA and Streck plasma samples containing native and crosslinked cf-nucleosomes, respectively, were collected from 10 asymptomatic healthy volunteers, 49 hospitalized patients with elevated levels of CRP >5 (INO Specimens Biobank) and 25 patients diagnosed with a solid cancer (Lyon).

Inclusion criteria for healthy subject samples were age ≥18 years and self-declared as not having any known disease diagnosis. Samples from healthy subjects aged <18 years were excluded. Inclusion criteria for cancer patient samples were age ≥18 years and a confirmed, treatment-naïve cancer diagnosis. Exclusion criteria for cancer patient samples were age <18 years or having previously received cancer treatment. Inclusion criteria for samples from hospitalized patients with elevated levels of CRP were hospitalization, age ≥18 years and a CRP level >5 ng/mL. Exclusion criteria for samples from hospitalized patients with elevated CRP were age <18 years or a CRP level ≤5 ng/mL. Details of age and sex of healthy subjects and cancer patients are provided in **Supplementary Tables S1** and **S2**.

Laboratory investigations were blinded, but the protocol was not double-blinded and sample collection was not randomized. As this was a first experimental investigation of the effect crosslinking plasma cf-nucleosomes, the protocol was not registered at ClinicalTrials.gov.

Informed consent was obtained from all subjects involved in the study and all experiments and methods were performed in accordance with relevant guidelines and regulations. The study followed the principles outlined in the Declaration of Helsinki for all human experimental investigations. All blood specimens were purchased from biobanks where the blood samples are ethically collected with Institutional Review Board-approved protocols. INO Specimens Biobank is authorized to market residual samples from medical analysis to carry out scientific research (file number: AC-2018-3151 and authorization number: IE-2018-978).

### Automated immunoassays

Assays for nucleosomes containing the histone isoform H3.1 (H3.1-nucleosomes) and for modified nucleosomes containing the histone modification H3K27Me3 (H3K27Me3-nucleosomes) or H3K36Me3 (H3K36Me3-nucleosomes) were performed in 50 µL of plasma using Nu.Q^®^ Immunoassays (manufactured by Belgian Volition SRL, Isnes, Belgium) with an automated random-access chemiluminescence immunoassay analyzer system (IDS-i10, manufactured by IDS Immunodiagnosticsystems). Results measured at Belgian Volition SRL were the mean of duplicate analysis. Results measured at Centre Hospitalier Lyon Sud were single measurements.

### Manual immunoassays

Manual 96-well photometric cf-nucleosome immunoassays were performed using the Cell Death Detection ELISA (Roche Diagnostics GmbH, Mannheim, Germany) and the Nu.Q^®^ Discover H3.1 ELISA Research Use Only Kit (Belgian Volition SRL, Belgium) according to manufacturers’ instructions. Results measured were the mean of duplicate analysis.

### Plasma cf-nucleosome ChIP-Seq

DNA fragment size frequency diagrams were produced for cfDNA extracted from 1 mL Streck plasma and from cf-nucleosomes immunoprecipitated from 1 mL Streck plasma. cf-nucleosome immunoprecipitation of samples collected from four treatment-naïve patients with CRC was performed by incubation of MyOneTosyl magnetic beads (Thermo Fisher Scientific, Waltham, MA, USA) coated with 30 µg/mg of anti-H3.1-nucleosome antibody for 1 hour at room temperature. The beads were isolated and washed five times with RIPA buffer. DNA was extracted from whole plasma and from the immunoprecipitated cf-nucleosomes using a QIAamp^®^ DSP Circulating NA kit from QIAGEN (61504 (IVD)). Extracted cfDNA was eluted in 50 µL and quantified using a Qubit dsDNA HS assay kit from Thermo Fisher (Q32854). 5 ng DNA was used for single-strand library preparation using the SRSLY PicoPlus Base Kit from ClaretBio (#CBS-K25B-24). DNA sequencing was performed using an Illumina NovaSeq 6000 Sequencing System.

Bioinformatic analysis was performed using the nf-core ATAC-seq/ChIP-seq pipeline v2.1.2, including adapter trimming (Trim Galore v.0.6.7, using 20 bp trimming to accommodate NovaSeq chemistry), alignment (BWA-MEM v.0.7.17-r1188), blacklist filtering, and low-quality read filtering (duplicate reads, non-primary and multi-mapping reads, and reads containing more than four mismatches or improperly paired).

Fragment size frequency diagrams were produced for filtered sequences using the CollectInsertSizeMetrics tool from Picard (v3.0.0), and plotted using Multi-QC v1.13.

Further details of the bioinformatic tools used are available from Patel et al (29), Ewels et al (30), and The Boyle Lab (31).

### Data availability

Details of the data generated in this study are available upon request from the corresponding author.

## Results

### Automated immunoassay results for crosslinked H3.1-nucleosomes

A first sample set collected from 100 asymptomatic healthy volunteers, 229 patients diagnosed with cancer, 10 non-hospitalized patients diagnosed with an inflammatory condition (sourced from BioIVT and Indivumed), and 49 hospitalized patients with elevated levels of the inflammatory marker CRP (sourced from INO Specimens) were analyzed at Belgian Volition SRL, Belgium (Volition). Details of samples assayed at Volition are provided in **Supplementary Tables S1** and **S2**. The results obtained are shown in **Fig. 1**. The range of results observed for healthy subjects aged 18–78 years was 2.3–31.7 ng/mL. No increase in levels of crosslinked cell-free H3.1-nucleosomes (cf-H3.1-nucleosomes) with age was observed (**Supplementary Fig. S1**). The range of results in non-hospitalized patients with an inflammatory disorder (not in a flare) was 2.3–29.4 ng/mL. The range of results in cancer patients was 3.6–>6000 ng/mL and levels were elevated compared with those of asymptomatic healthy controls (Wilcoxon-Mann-Whitney p-value = 5 x 10^-24^). Consistent with previous observations that cf-nucleosome levels are raised in a variety of non-cancer conditions, particularly inflammatory conditions (5), we observed cf-nucleosome levels in the range 11.2–591.8 ng/mL in hospitalized patients with elevated levels of the inflammatory marker CRP.

**Figure 1.**
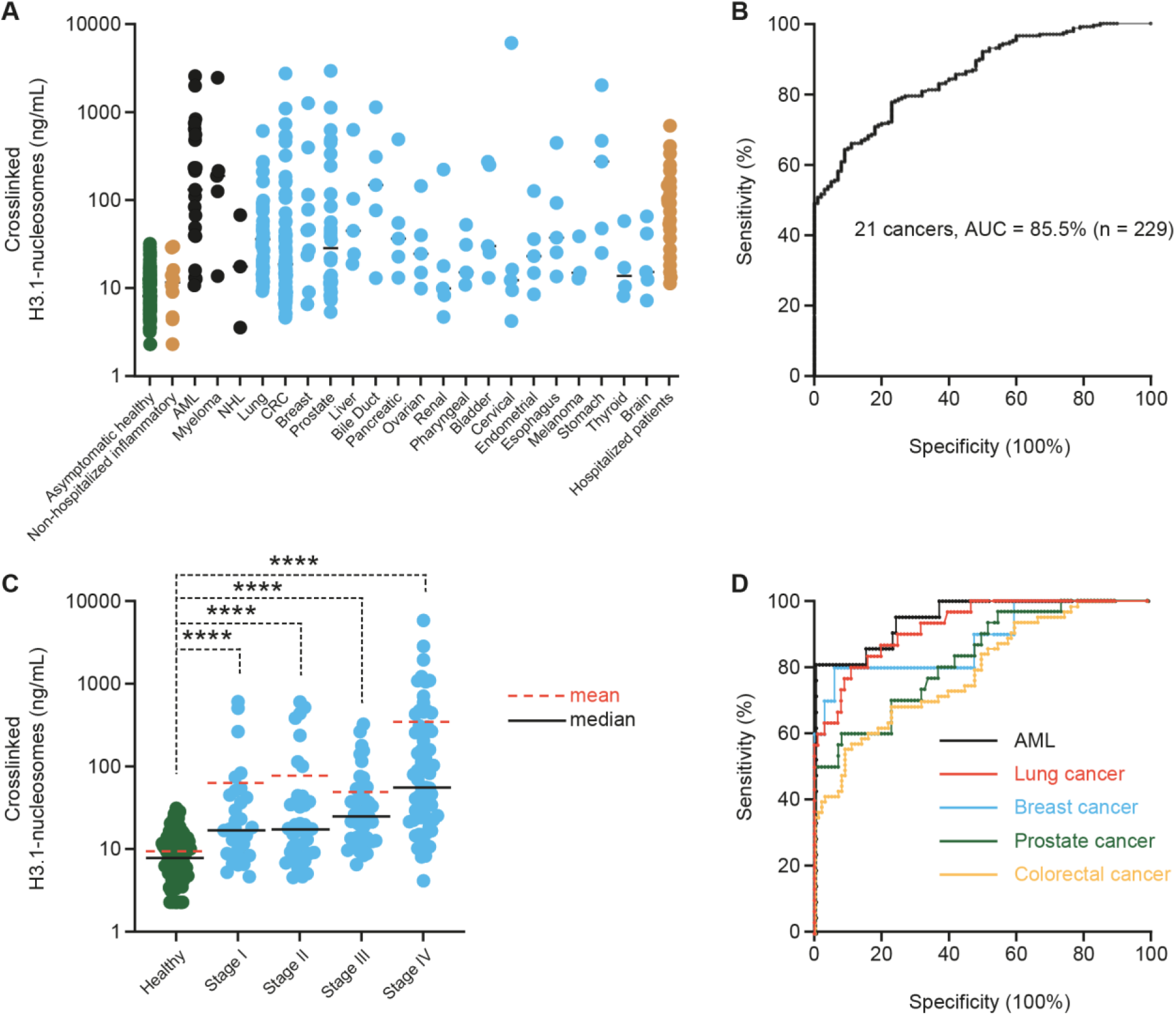
**A**, Dot plot of H3.1-nucleosome levels measured in plasma samples from asymptomatic healthy volunteers (green, n = 100), non-hospitalized patients diagnosed with an inflammatory disease (beige, n = 10), a liquid cancer (black, n = 29), a solid cancer (blue, n = 200) or hospitalized patients with an elevated level CRP (beige, n=49). **B,** ROC curve for detection of 229 samples representing 21 cancer types (control n = 100). **C,** Dot plot of H3.1-nucleosome levels measured in plasma samples from patients diagnosed with a solid cancer by stage (blue; n = 177); asymptomatic healthy control: median result = 8.1 ng/mL (range 2.3–31.7), Stage I: 17.4 (4.7–622), Stage II: 17.9 (4.6–620), Stage III: 25.3 (6.6–332), Stage IV: 57.2 (4.2–>6000). **D,** ROC curves for detection of some common cancers (control n = 100).

As high specificity is important in screening tests to minimize false-positive results, we set a cutoff value for a positive result in cancer patients as greater than the highest value observed for any asymptomatic healthy control sample (>31.7 ng/mL). Using this cutoff, the results for 112 of 229 cancer patients were positive, giving an overall sensitivity of 49% for cancer detection in this sample set, with zero false-positive results among our cohorts of asymptomatic healthy subjects (**Fig. 1**). The results for 10 subjects diagnosed with an inflammatory disorder were all negative (<31.7 ng/mL) **(Fig. 1A)**. Receiver operator characteristic (ROC) curves for all 229 cancer patients (**Fig. 1B**) and for some common cancers (**Fig. 1D**) are shown at 100% observed specificity (control n = 100).

We investigated the dependency of crosslinked cf-nucleosome levels on cancer stage in 177 of the 229 cancer patients who were diagnosed with a solid cancer of known disease stage. Levels of crosslinked cf-nucleosomes were elevated over those observed in asymptomatic healthy volunteers for some solid cancer patients diagnosed at all disease stages (Wilcoxon-Mann-Whitney p-values = 8 x 10^-6^, 7 x 10^-7^, 4 x 10^-12^ and 5 x 10^-20^ for cancer stages I, II, III and IV, respectively). The sensitivity of the automated H3.1-nucleosome assay for detection of each disease stage with zero false-positive results among the asymptomatic control cohorts is shown for all 177 subjects with a solid cancer, and for some common individual cancers, in **Table 1** and **Fig. 1C**.

**Table 1.** Stage-dependency for sensitivity (positive results/number of samples tested) for solid cancer detection at 100% observed specificity (control n = 100).

| Stage | Lung | Liver and bile duct | Breast | Prostate | CRC | All solid cancers (n = 177) |
| --- | --- | --- | --- | --- | --- | --- |
| I | 29% (2/7) | 100% (2/2) | 33% (1/3) | 50% (3/6) | 10% (1/10) | 34% (11/32) |
| II | 38% (3/8) | 100% (2/2) | 50% (1/2) | 63% (5/8) | 12% (2/17) | 39% (15/38) |
| III | 67% (4/6) | 0% (0/2) | 50% (1/2) | 29% (2/7) | 38% (6/16) | 42% (18/43) |
| IV | 89% (8/9) | 75% (3/4) | 100% (3/3) | 56% (5/9) | 71% (12/17) | 67% (43/64) |

A second independent sample set collected from 50 asymptomatic healthy volunteers and 160 patients diagnosed with cancer (sourced from Etablissement Français du Sang and from Centre Hospitalier Lyon Sud, respectively) was analyzed for H3.1-nucleosome levels at Lyon. The range of crosslinked H3.1-nucleosome levels measured in Streck plasma samples from 50 asymptomatic healthy blood donors was 1.8–19.1 ng/mL. The range of results in 160 CRC and lung cancer patients was 2.2–>6000 ng/mL. Comparative dot plots and ROC curves obtained by analysis of different (non-matched) sample cohorts at Volition and Lyon are shown in **Fig. 2**.

**Figure 2.**
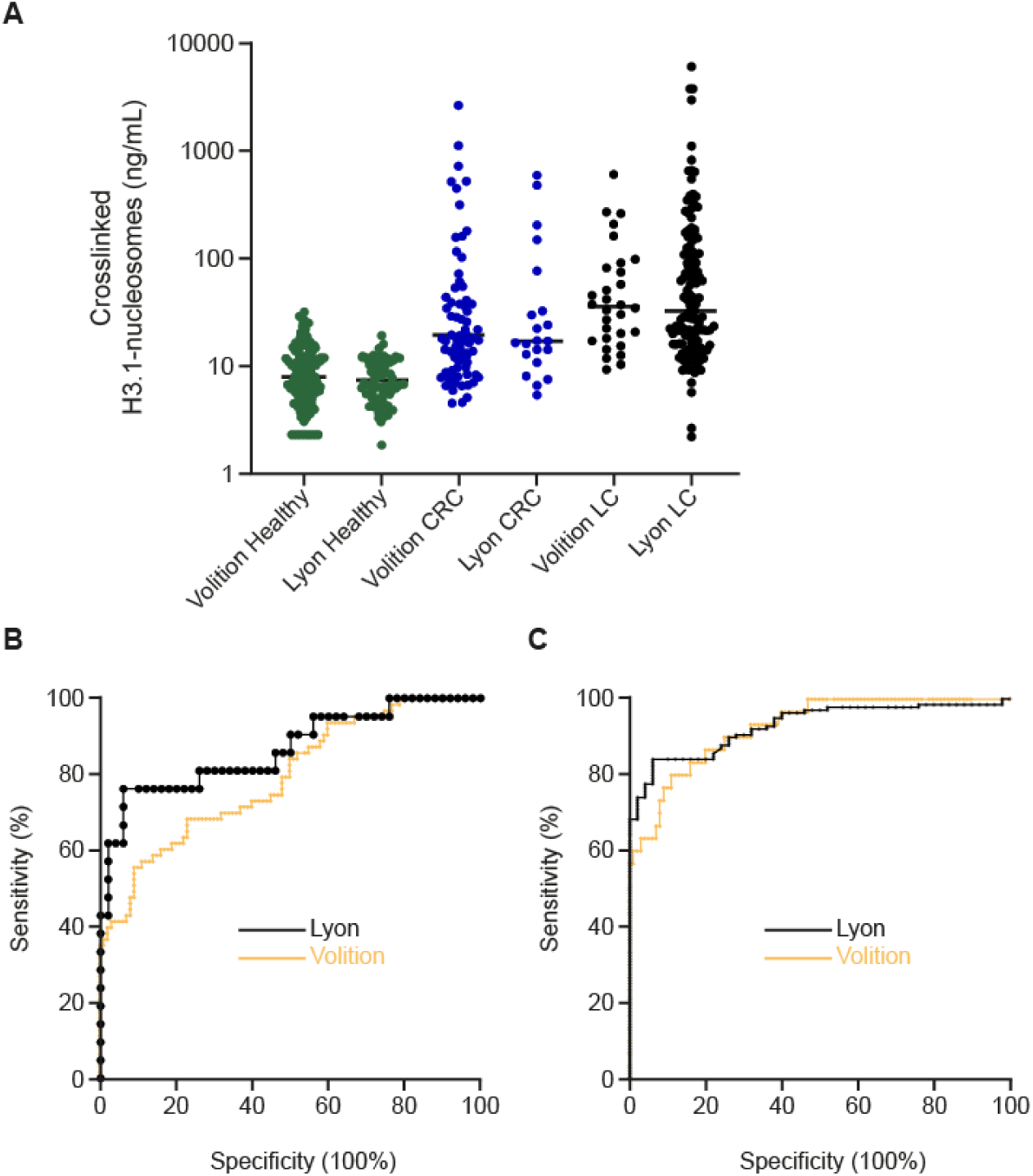
**A**, Dot plot showing crosslinked H3.1-nucleosome levels observed for independent asymptomatic healthy and diseased (colorectal cancer [CRC] or lung cancer [LC]) subject cohorts analyzed independently at Lyon or Volition. No significant difference was observed between the results for healthy subjects or for patients diagnosed with LC or CRC obtained at Lyon or Volition (Wilcoxon-Mann-Whitney p-value = 0.3601, 0.9852 or ≈1.0, respectively). **B,** ROC curves obtained by automated immunoassay of crosslinked H3.1-nucleosomes by Volition and Lyon for CRC (Volition CRC: AUC = 79.0%, n = 63 [46 Stage I–III, 13 Stage IV, four unknown stage]; Lyon CRC: AUC = 86.7%, n = 21 [seven Stage I–III, 14 Stage IV]). **C,** ROC curves for LC (Volition LC: AUC = 92.5%, n = 30 [21 Stage I–III, nine Stage IV]; Lyon LC: AUC = 93.0%, n = 139 [0 Stage I–II, 139 Stage III–IV]).

### Automated assay for nucleosomes containing post-translational modifications

We investigated whether the results obtained for crosslinked cf-nucleosomes were exclusive to the automated H3.1-nucleosome assay or also occur for other cf-nucleosome structure assays. We measured the levels of crosslinked H3K27Me3- and H3K36Me3-nucleosomes in a 166-patient subset of the 339 BioIVT and Indivumed patient cohort described above (47 asymptomatic healthy subjects and 119 subjects with a cancer diagnosis) by automated chemiluminescence immunoassay. The two assays for modified nucleosomes produced results with similar cancer detection to the automated H3.1-nucleosome assay for the same samples (**Fig. 3A**).

**Figure 3.**
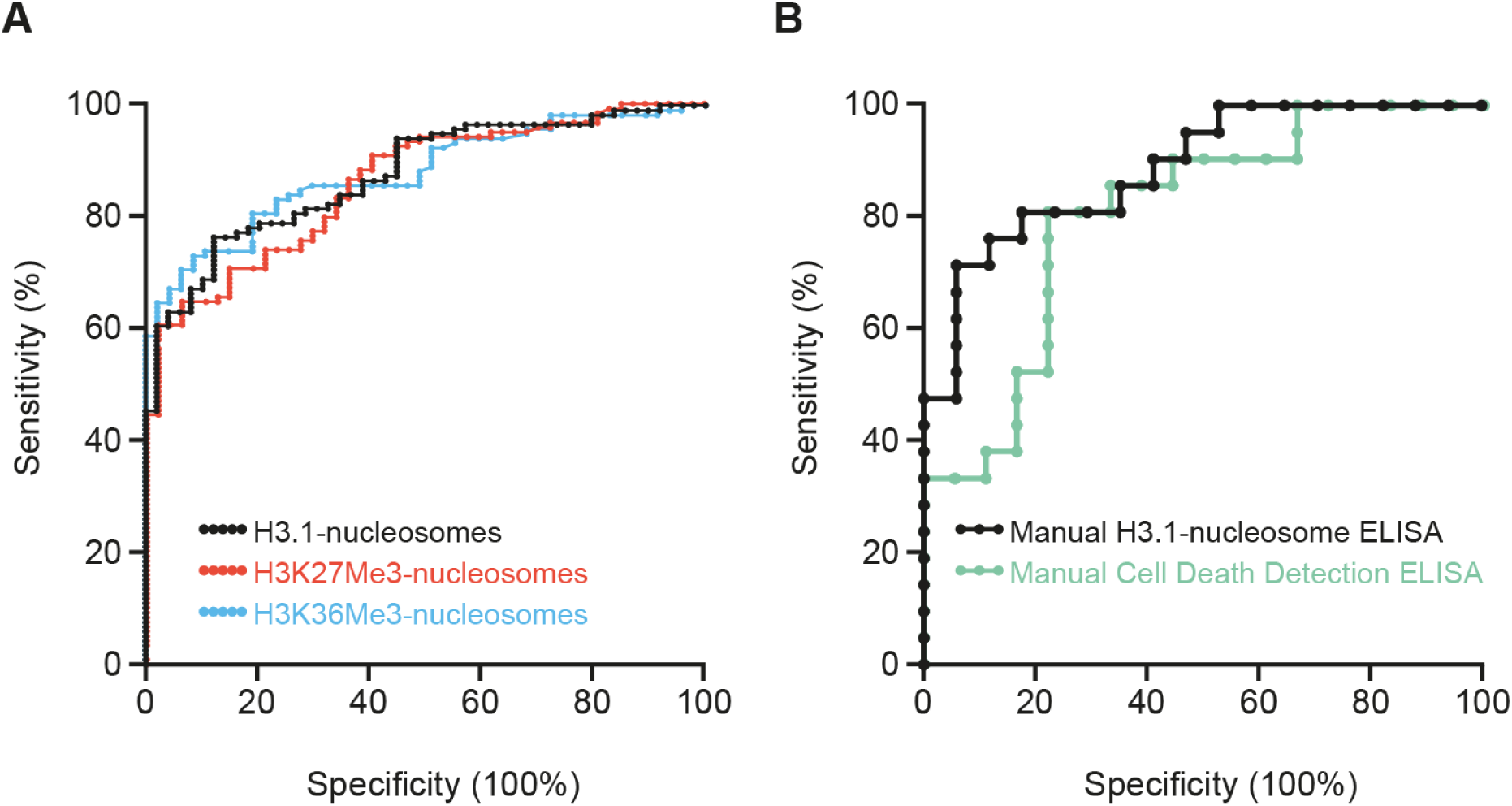
ROC curves generated for: **A,** Automated immunoassays for nucleosomes containing histone post-translational modifications at H3K27Me3 (AUC = 86.0%) or H3K36Me3 (AUC = 87.9%) and H3.1-nucleosomes for the same samples (AUC = 87.6%). **B,** Manual H3.1 ELISA kit (AUC = 88.8%) and Cell Death Detection ELISA kit (AUC = 80.7%) for the same samples.

### Manual immunoassays for cf-nucleosomes

To investigate whether the results obtained for the automated H3.1-nucleosome assay were reproducible using manual ELISA kits, we measured the level of crosslinked H3.1-nucleosomes in a 39-patient subset (18 asymptomatic healthy subjects and 21 subjects with CRC) of the 339-patient cohort described above using the manual Volition Nu.Q^®^ Discover H3.1 ELISA and the manual Roche Cell Death Detection ELISA. Both manual ELISA kits detected CRC but we observed higher sensitivity at 100% specificity (48% vs 33%) and higher AUC (89% vs 81%) for the Nu.Q^®^ Discover kit than the Cell Death Detection ELISA (**Fig. 3B**).

### Effect of crosslinking nucleosomes in whole blood

The effect of crosslinking cf-nucleosomes in whole blood samples (prior to sample processing) on immunoassay results was investigated by comparing results obtained for matched EDTA plasma (native) and Streck plasma (crosslinked) samples collected from 10 asymptomatic healthy volunteers, 49 hospitalized patients with elevated levels of CRP and 25 patients diagnosed with a solid cancer. We observed that crosslinking cf-nucleosomes in whole blood by sample collection in Streck Cell-Free DNA BCTs led to an increase in measured cf-nucleosome level in 10 of 25 (40%) cancer samples tested (mean increase +23%, range -76% to +343%), but in zero of 59 samples from asymptomatic healthy volunteers or hospitalized patients with elevated levels of CRP (**Fig. 4 and Supplementary Table S3a**).

**Figure 4.**
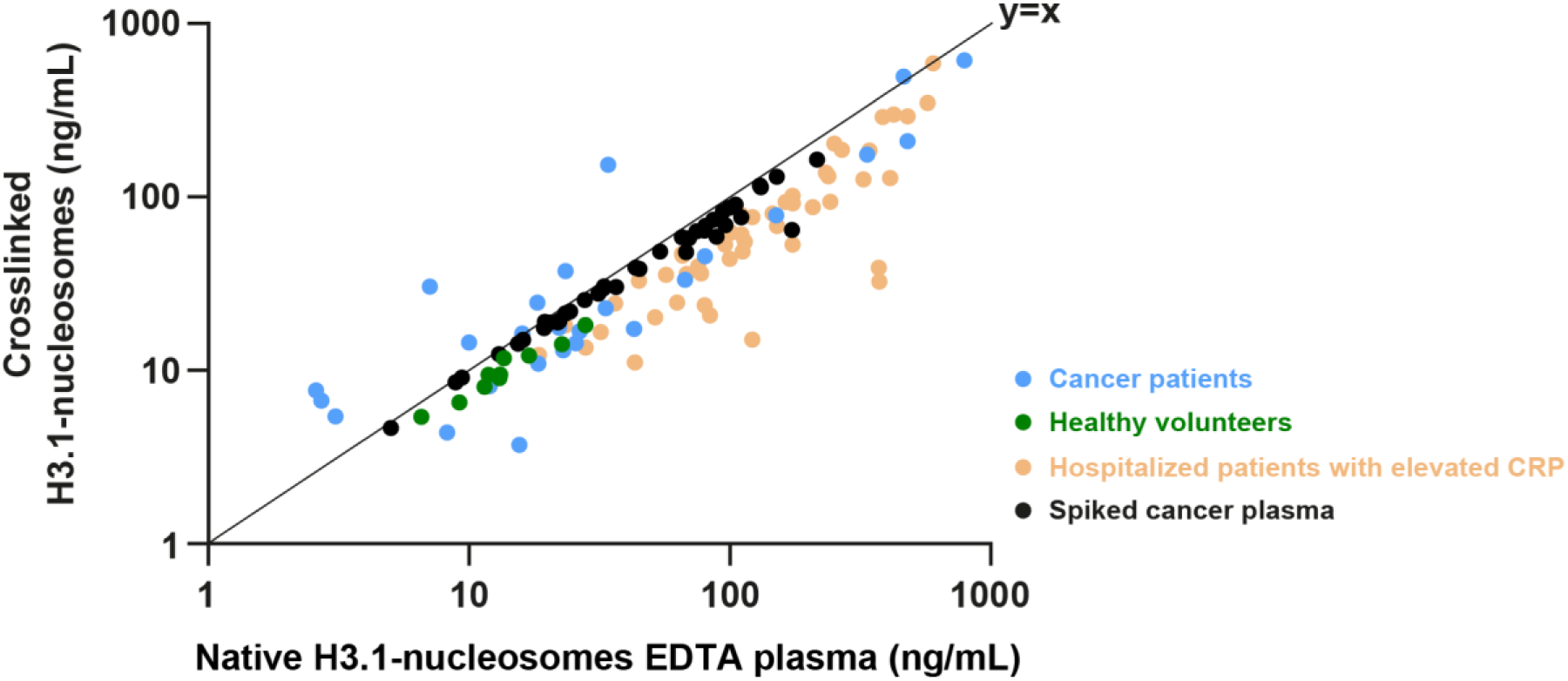
H3.1-nucleosome levels measured after crosslinking in whole blood by sample collection in EDTA BCTs (native) or Streck Cell-Free DNA BCTs (crosslinked) obtained from; **Green**, 10 asymptomatic healthy volunteers. **Light brown**, 49 hospitalized patients with elevated levels of CRP (>5 mg/L). **Blue**, 25 patients diagnosed with a solid cancer. **Black**, H3.1-nucleosome levels measured after crosslinking in EDTA plasma by addition of 3.33% v/v assay buffer (native) or 3.33% Streck BCT additive (crosslinked) to EDTA plasma samples obtained from 40 patients with CRC.

Among samples from healthy subjects, we observed a mean decrease in measured cf-nucleosome results of -27% (range -38% to -14%) on crosslinking. Among samples from hospitalized patients with elevated levels of CRP, we observed a mean decrease in measured results of -47% (range -91% to -1%) on crosslinking. In a minority of samples, particularly samples from hospitalized patients with elevated levels of CRP, crosslinking led to a large reduction in measured cf-nucleosome levels of up to -91% (**Fig. 4** **and Supplementary Table S3a**).

### Effect of crosslinking cf-nucleosomes in EDTA plasma

We next investigated whether the observed increase in measured cf-nucleosome levels in some cancer patients was exclusive to crosslinking in whole blood samples, or would also occur for cf-nucleosomes crosslinked after sample processing in EDTA plasma.

EDTA plasma samples from 40 patients diagnosed with CRC were spiked with 3.33% v/v Streck BCT additive (crosslinked) or buffer (native), incubated for 1 hour at room temperature and assayed for H3.1-nucleosomes using the automated immunoassay.

Crosslinking by spiking plasma with Streck additive led to a lower measured cf-nucleosome result in all 40 samples, with a mean decrease of -14% (range -62% to -2%) on crosslinking, demonstrating that an increased level occurs only for samples crosslinked in whole blood and not for cf-nucleosomes crosslinked in processed plasma (**Fig. 4** **and Supplementary Table S3b**).

### Plasma cf-nucleosome ChIP-Seq

Intact plasma cf-mononucleosomes comprise a histone core and approximately 130– 200 bp DNA. Nicked nucleosomes, in which the cf-nucleosome-associated DNA fragment includes one or more single-or double-stranded cuts or nicks, would be expected to yield shorter DNA fragments. To ascertain whether any short fragments occur in crosslinked cf-nucleosomes from cancer patients, and whether these fragments are bound by the solid phase antibody employed in the H3.1-nucleosome immunoassay, we investigated the fragment size frequency profile for cfDNA associated with crosslinked H3.1-nucleosomes bound by the antibody.

cf-nucleosomes were isolated by ChIP from Streck plasma samples collected from four patients with CRC using the magnetic solid phase antibody employed in the H3.1-nucleosome immunoassay. DNA extracted from the antibody-bound crosslinked cf-nucleosomes was sequenced and fragment size frequency profiles produced. Consistent with previous findings (17–19), the profiles confirm the presence of a significant, but variable, level of nicked, crosslinked cf-nucleosome-derived short cfDNA fragments in the four CRC patient samples. The results also indicate that crosslinked, nicked cf-nucleosomes are bound by the magnetic antibody used in the H3.1-nucleosome assay (**Fig. 5**). DNA fragment size profiles obtained for the ChIP isolates were similar to profiles for cfDNA extracted directly from the same whole Streck plasma samples, confirming that most plasma cfDNA is nucleosome-protected and indicating a high efficiency of immunoprecipitation (**Supplemental Fig. S2**).

**Figure 5.**
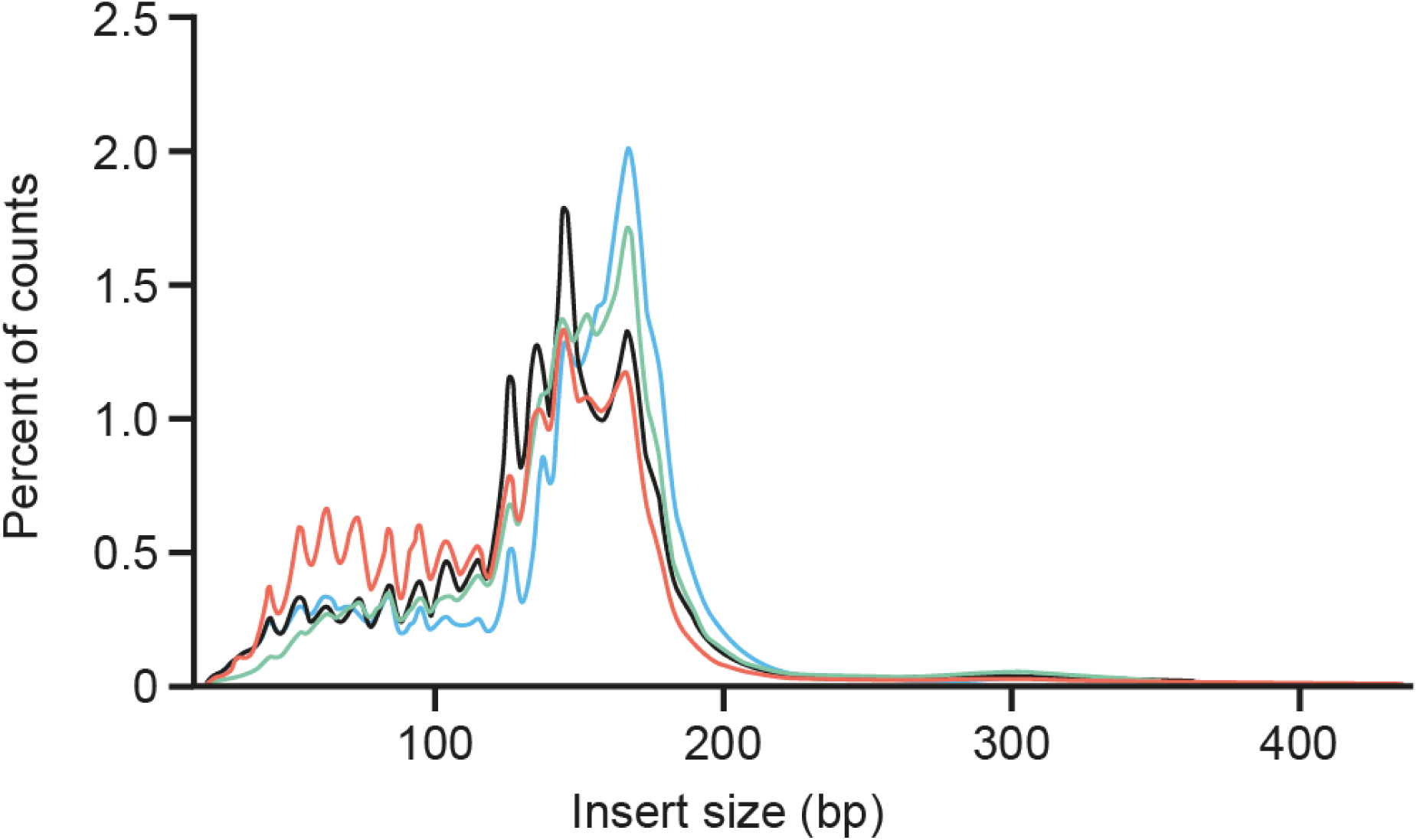
cfDNA fragment size frequency profiles for cf-nucleosome-associated cfDNA isolated by anti-H3.1-nucleosome ChIP from Streck plasma samples collected from four patients with CRC.

## Discussion

Current population-based cancer screening methods among asymptomatic subjects are not specific for cancer. A majority of positive screening results obtained by LDCT, mammography, FIT and MRI testing for lung, breast, CRC or prostate cancer, respectively, are false-positive results as follow-up investigations find no cancer (32–35). Plasma biomarkers to aid in the interpretation of positive screening results may accelerate diagnosis and treatment for cancer patients, reduce unnecessary invasive follow-up investigations, and reduce costs.

To minimize false-positive results in the current experiments, we set a cutoff at the highest level of crosslinked plasma cf-H3.1-nucleosomes observed for any control subject in the first 100 subjects tested of the asymptomatic healthy cohort. All results obtained for a second, independently collected and tested, 50-subject asymptomatic cohort were below the level of the cutoff selected. Using the cutoff, we observed elevated levels of crosslinked plasma cf-H3.1-nucleosomes in approximately half of all newly diagnosed treatment-naïve cancer patients tested, including a third of patients with Stage I disease. An elevated plasma level of crosslinked cf-nucleosomes is a non-specific finding associated with a variety of pathological conditions, and is not diagnostic of cancer. However, our results show that an elevated level of crosslinked cf-H3.1-nucleosomes in an asymptomatic subject is indicative of an increased likelihood of cancer. The positive signal we observed for 34% and 39% of Stage I and II cancers, respectively, with no false-positive results in the combined 150-subject asymptomatic cohorts studied, suggests cf-nucleosome measurements may have utility to identify asymptomatic subjects at high risk for cancer to aid the interpretation of positive screening results.

The potential for crosslinked cf-nucleosome measurements was not restricted to a particular nucleosome assay; similar findings were observed for a number of automated or manual nucleosome assays.

Nicked cf-nucleosomes are reported to be more prevalent in plasma samples from cancer patients than healthy subjects (17–19). Consistent with these reports, we observed that nucleosome-derived sub-100 bp cfDNA fragments accounted for a significant proportion of the cfDNA present in the plasma of four CRC patients investigated. Interestingly, sub-100 bp cfDNA fragments are not detectable by next-generation sequencing methods employing conventional double-stranded library preparation, but may be analyzed using single-stranded DNA library preparation (36, 37).

Clipped cf-nucleosomes may be involved in cancer development and proliferation. Metalloproteases responsible for clipping the tails of histones H2, H3, or H4 are highly expressed in cancer patients and critical for proliferation of cancer cells. Depletion of metalloproteases greatly decreases cell proliferation in breast cancer cells (15).

Nicked and clipped cf-nucleosomes are reported to be less stable than intact cf-nucleosomes and require Ca^2+^ and Mg^2+^ to maintain conformation (10–12, 14, 15). Crosslinking is commonly used to stabilize nucleoproteins in cellular ChIP-seq studies but has not previously been reported in immunoassay of plasma cf-nucleosomes. We compared head-to-head cf-nucleosome results in matched pairs of native EDTA and crosslinked Streck plasma samples from cancer patients, asymptomatic healthy volunteers and hospitalized patients with elevated inflammatory markers. Crosslinking of cf-nucleosomes in whole blood led to an increase in measured cf-nucleosome level in 10 of 25 cancer samples, but to a reduction in every non-cancer sample tested, whether healthy or diseased. This increase only occurred where the crosslinking was performed in whole blood and did not occur when cf-nucleosomes were crosslinked in pre-processed EDTA plasma (**Fig. 4** **and Supplementary Table S3**). The chemical mechanism underlying these findings is unclear and requires further research, but the results are consistent with a hypothesis that less stable nicked or clipped cf-nucleosomes are more prevalent in cancer than healthy plasma and that their conformation may be conserved by crosslinking immediately at venipuncture, thereby preserving their structure for antibody binding in immunoassay.

Formaldehyde-releasing agents are used in cfDNA BCT to stabilize cell membranes by crosslinking and to prevent cell lysis and contamination of plasma with intracellular genomic DNA. In addition, formaldehyde or formaldehyde-releasing agents are reported to crosslink other proteins more widely (27), and to crosslink csb-cf-nucleosomes to cell membranes (22, 23). In a minority of samples, particularly samples from hospitalized patients with elevated levels of CRP, we observed that crosslinking in whole blood led to a large reduction (up to 91%) in measured cf-nucleosome levels (**Fig. 4** **and Supplementary Table S3**). The mechanism underlying this finding may relate solely to the intended purpose of Streck Cell-Free DNA BCTs in preventing the release of cellular DNA into plasma, but may also in part be associated with crosslinking of csb-cf-nucleosomes to cell membranes in whole blood collected in Streck tubes, thereby preventing elution into plasma as described by Bryzgunova (20, 21). This is an area for further research.

This study was designed as a first experimental investigation into the feasibility of crosslinked plasma cf-nucleosome measurements by immunoassay and their potential utility as a marker for cancer in asymptomatic subjects. Further research is needed to fully understand the underlying mechanisms and evaluate potential clinical utility. The present study has a number of limitations including: (i) the healthy volunteer cohort investigated may not be representative of asymptomatic screening populations, (ii) the cancer cohort was not a screen-diagnosed population, (iii) the sample numbers of the individual cancer types studied were low, (iv) no evaluation of minimal residual disease, or prognostication for potential utility in disease management was made, (v) cf-nucleosome results were not investigated in cancer screen-positive and screen-negative patients, and (vi) further mechanistic investigations into the chemistry of cf-nucleosomes in plasma are required.

In summary, our study demonstrates that immunoassay measurement of crosslinked plasma cf-nucleosomes is a novel promising minimally invasive approach to aid in the early detection of cancer among asymptomatic individuals. The consistency of results across different testing sites, assay platforms, and nucleosome species provides robust technical validation of the approach. The method achieved clinically meaningful sensitivity (49% overall, 34% in Stage I disease) at 100% observed specificity among asymptomatic subjects in the cohorts studied. This compares favorably with existing ctDNA-based early detection approaches (38). The ability to detect early-stage cancers across multiple tumor types is of particular interest, suggesting the potential for broad utility as an adjunct to current screening methods.

## Authors’ Contributions

Conceptualization: **J. Micallef** and **D. Pamart**; Methodology: **J. Micallef**, **D. Pamart**, **B. Cuvelier** and **M. Herzog**; Investigation: **D. Pamart**, **G. Rommelaere**, **B. Cuvelier**, **A. Govaerts**, **L. Payen** and **M. Piecyk**; Formal Analysis: **J. Micallef** and **D. Pamart**; Visualization: **J. Micallef**; Writing – Original Draft: **J. Micallef**; Writing – Review & Editing: **J. Micallef**; **D. Pamart**, **M. Herzog**, **G. Rommelaere**, **L. Payen** and **M. Piecyk**; Data Curation: **D. Pamart** and **J. Micallef**; Funding Acquisition: **M. Herzog**; Supervision: **J. Micallef**, **D. Pamart** and **M. Herzog**.

## Acknowledgments

We thank the National University of Taiwan for EDTA plasma samples; Ben Berman, Daniel Halter and Jacques Avaux for bioinformatics; Monika Suchora of BioIVT for timed blood sample collection; Andrew Retter for review and advice in manuscript preparation; and Thomas Bygott for statistical advice. Graphical design and development by Jim Park (Sparked into Life, Macclesfield, UK). Medical writing support, including development of drafts in consultation with the authors, collating author comments, copyediting and fact checking was provided by Rachel Mason (Macclesfield, UK). Funding was provided by Belgian Volition SRL. We thank AstraZeneca for their financial support of the CIRCAN program to establish liquid biopsy in routine care.

## References

1. Moleyar-Narayana P, Leslie SW, Ranganathan S. Cancer screening. StatPearls: StatPearls Publishing. Copyright © 2025, StatPearls Publishing LLC.; 2025.

2. Wilson-Robles HM, Bygott T, Kelly TK, Miller TM, Miller P, Matsushita M, et al. Evaluation of plasma nucleosome concentrations in dogs with a variety of common cancers and in healthy dogs. BMC Vet Res 2022;18:329.

3. Wilson-Robles H, Warry E, Miller T, Jarvis J, Matsushita M, Miller P, et al. Monitoring plasma nucleosome concentrations to measure disease response and progression in dogs with hematopoietic malignancies. PLoS One 2023;18:e0281796.

4. Grolleau E, Candiracci J, Lescuyer G, Barthelemy D, Benzerdjeb N, Haon C, et al. Circulating H3K27 methylated nucleosome plasma concentration: synergistic information with circulating tumor DNA molecular profiling. Biomolecules 2023;13:1255.

5. Wang H, Wang Y, Zhang D, Li P. Circulating nucleosomes as potential biomarkers for cancer diagnosis and treatment monitoring. Int J Biol Macromol 2024;262:130005.

6. Garrido MM, Ribeiro RM, Krüger K, Pinheiro LC, Guimarães JT, Holdenrieder S. Relevance of circulating nucleosomes, HMGB1 and sRAGE for prostate cancer diagnosis. In Vivo 2021;35:2207–12.

7. Wiig H, Kolmannskog O, Tenstad O, Bert JL. Effect of charge on interstitial distribution of albumin in rat dermis in vitro. J Physiol 2003;550:505–14.

8. Korolev N, Lyubartsev AP, Nordenskiöld L. A systematic analysis of nucleosome core particle and nucleosome-nucleosome stacking structure. Sci Rep 2018;8:1543.

9. Vengerov YY, Popenko VI. Changes in chromatin structure induced by EDTA treatment and partial removal of histone H1. Nucleic Acids Res 1977;4:3017–27.

10. Yang Z, Hayes JJ. The divalent cations Ca2+ and Mg2+ play specific roles in stabilizing histone-DNA interactions within nucleosomes that are partially redundant with the core histone tail domains. Biochemistry 2011;50:9973–81.

11. Ohyama T. New aspects of magnesium function: a key regulator in nucleosome self-assembly, chromatin folding and phase separation. Int J Mol Sci 2019;20.

12. Strick R, Strissel PL, Gavrilov K, Levi-Setti R. Cation-chromatin binding as shown by ion microscopy is essential for the structural integrity of chromosomes. J Cell Biol 2001;155:899–910.

13. Gebala M, Johnson SL, Narlikar GJ, Herschlag D. Ion counting demonstrates a high electrostatic field generated by the nucleosome. Elife 2019;8:e44993.

14. Morioka S, Oishi T, Hatazawa S, Kakuta T, Ogoshi T, Umeda K, et al. High-speed atomic force microscopy reveals the nucleosome sliding and DNA unwrapping/wrapping dynamics of tail-less nucleosomes. Nano Lett 2024;24:5246–54.

15. Liu H, Wang C, Lee S, Deng Y, Wither M, Oh S, et al. Clipping of arginine-methylated histone tails by JMJD5 and JMJD7. Proc Natl Acad Sci U S A 2017;114:E7717–e26.

16. Imre L, Simándi Z, Horváth A, Fenyőfalvi G, Nánási P, Niaki EF, et al. Nucleosome stability measured in situ by automated quantitative imaging. Sci Rep 2017;7:12734.

17. Thierry AR. Circulating DNA fragmentomics and cancer screening. Cell Genom 2023;3:100242.

18. Mouliere F, Robert B, Arnau Peyrotte E, Del Rio M, Ychou M, Molina F, et al. High fragmentation characterizes tumour-derived circulating DNA. PLoS One 2011;6:e23418.

19. Andersen RF, Spindler KL, Brandslund I, Jakobsen A, Pallisgaard N. Improved sensitivity of circulating tumor DNA measurement using short PCR amplicons. Clin Chim Acta 2015;439:97–101.

20. Bryzgunova OE, Tamkovich SN, Cherepanova AV, Yarmoshchuk SV, Permyakova VI, Anykeeva OY, et al. Redistribution of free- and cell-surface-bound DNA in blood of benign and malignant prostate tumor patients. Acta Naturae 2015;7:115–8.

21. Tamkovich S, Laktionov P. Cell-surface-bound circulating DNA in the blood: biology and clinical application. IUBMB Life 2019;71:1201–10.

22. Wang H, Shan X, Ren M, Shang M, Zhou C. Nucleosomes enter cells by clathrin- and caveolin-dependent endocytosis. Nucleic Acids Res 2021;49:12306–19.

23. Watson K, Gooderham NJ, Davies DS, Edwards RJ. Nucleosomes bind to cell surface proteoglycans. J Biol Chem 1999;274:21707–13.

24. Hoffman EA, Frey BL, Smith LM, Auble DT. Formaldehyde crosslinking: a tool for the study of chromatin complexes. J Biol Chem 2015;290:26404–11.

25. Gavrilov A, Razin SV, Cavalli G. In vivo formaldehyde cross-linking: it is time for black box analysis. Brief Funct Genomics 2015;14:163–5.

26. Jackson V. Formaldehyde cross-linking for studying nucleosomal dynamics. Methods 1999;17:125–39.

27. Röth D, Molina-Franky J, Williams JC, Kalkum M. Mass spectrometric detection of formaldehyde-crosslinked PBMC proteins in cell-free DNA blood collection tubes. Molecules 2023;28:7880.

28. Poorey K, Viswanathan R, Carver MN, Karpova TS, Cirimotich SM, McNally JG, et al. Measuring chromatin interaction dynamics on the second time scale at single-copy genes. Science 2013;342:369–72.

29. Patel H, Espinosa-Carrasco J, Langer B, Ewels P, Garcia MU, Syme R, et al. nf-core/atacseq: [2.1.2] - 2022-08-07. 2023. Available from: https://zenodo.org/records/8222875.

30. Ewels P, Magnusson M, Lundin S, Käller M. MultiQC: summarize analysis results for multiple tools and samples in a single report. Bioinformatics 2016;32:3047–8.

31. The Boyle Lab. Blacklist / lists / hg38-blacklist.v2.bed.gz. Available from: https://github.com/Boyle-Lab/Blacklist/blob/master/lists/hg38-blacklist.v2.bed.gz.

32. Hammer MM, Byrne SC, Kong CY. Factors influencing the false positive rate in CT lung cancer screening. Acad Radiol 2022;29 Suppl 2:S18–s22.

33. Destounis S, Arieno A, Morgan R. New York State Breast Density Mandate: follow-up data with screening sonography. J Ultrasound Med 2017;36:2511–7.

34. Butterly LF, Hisey WM, Robinson CM, Limburg PJ, Kneedler BL, Anderson JC. What do ’false-positive’ stool tests really mean? Data from the New Hampshire Colonoscopy Registry. Prev Med Rep 2023;35:102309.

35. Eldred-Evans D, Burak P, Connor MJ, Day E, Evans M, Fiorentino F, et al. Population-based prostate cancer screening with magnetic resonance imaging or ultrasonography: the IP1-PROSTAGRAM study. JAMA Oncol 2021;7:395–402.

36. Snyder MW, Kircher M, Hill AJ, Daza RM, Shendure J. Cell-free DNA comprises an in vivo nucleosome footprint that informs its tissues-of-origin. Cell 2016;164:57–68.

37. Wang F, Li X, Li M, Liu W, Lu L, Li Y, et al. Ultra-short cell-free DNA fragments enhance cancer early detection in a multi-analyte blood test combining mutation, protein and fragmentomics. Clin Chem Lab Med 2024;62:168–77.

38. Bittla P, Kaur S, Sojitra V, Zahra A, Hutchinson J, Folawemi O, et al. Exploring circulating tumor DNA (CtDNA) and its role in early detection of cancer: a systematic review. Cureus 2023;15:e45784.

