## Supplementary tables for "Early Cancer Detection in Asymptomatic Subjects through Measurement of Crosslinked cf-Nucleosomes in Plasma"

**Supplementary Table S1.** Details of 100 Streck plasma samples collected from asymptomatic healthy volunteers and analyzed at Volition.

| Cohort | Sex (female/male) | Median age (range), years | Ethnicity |
|---|---|---|---|
| Indivumed (n = 20) | 10/10 | 59 (48–78) | African American (n = 16)<br>White Caucasian (n = 2)<br>Unknown (n = 2) |
| BioIVT (n = 80) | 45/25<br>Unknown (n = 10) | 31 (18–64)<br>Unknown (n = 10) | Asian (n = 7)<br>Black African (n = 6)<br>Black Caribbean (n = 1)<br>White Caucasian (n = 48)<br>White other (n = 2)<br>Other/Unknown (n = 16) |

**Supplementary Table S2**. Details of Streck plasma samples collected from 229 cancer patients and 10 patients with inflammatory disease analyzed at Volition by disease type. The ethnicity of subjects was unknown.

| **Disease source** | **Sex (female/male)** | **Median age (range), years** | **Disease stage** |
|---|---|---|---|
| Inflammatory disease (n = 10)<br>BioIVT | 8/2 | 36 (19–58) | N/A |
| AML (n = 21)<br>Indivumed | 14/7 | 77 (55–85) | N/A |
| Myeloma (n = 5)<br>Indivumed | 1/4 | 77 (70–87) | N/A |
| NHL (n = 3)<br>Indivumed | 2/1 | 59 (49–62) | Stage I: 1<br>Stage II: 1<br>Stage III:<br>Stage IV:<br>Stage X:<br>Unknown: 1 |
| Bladder (n = 5)<br>Indivumed | 0/5 | 73 (61–86) | Stage I:<br>Stage II:<br>Stage III: 2<br>Stage IV: 3<br>Stage X: |
| Brain (n = 5)<br>Indivumed | 4/1 | 74 (52–83) | Stage I:<br>Stage II:<br>Stage III:<br>Stage IV:<br>Stage X:<br>Unknown: 5 |
| Breast (n = 10)<br>Indivumed | 10/0 | 57 (43–78) | Stage I: 3<br>Stage II: 2<br>Stage III: 2<br>Stage IV: 3<br>Stage X: |
| Cervical (n = 5)<br>Indivumed | 5/0 | 66 (57–71) | Stage I:<br>Stage II:<br>Stage III: 1<br>Stage IV: 3<br>Stage X: 1 |
| CRC (n = 63)<br>Indivumed (n = 33)<br>BioIVT (n = 30) | 31/32 | 66 (50–82) | Stage I: 10<br>Stage II: 17<br>Stage III: 17<br>Stage IV: 16<br>Stage X: 3 |
| Endometrial (n = 5)<br>Indivumed | 5/0 | 68 (53–79) | Stage I:<br>Stage II:<br>Stage III: 1<br>Stage IV: 4<br>Stage X: |
| Esophageal (n = 5)<br>Indivumed | 0/5 | 70 (62–81) | Stage I:<br>Stage II: 1<br>Stage III: 1<br>Stage IV: 1<br>Stage X: 2 |
| Bile duct (n = 5)<br>Indivumed | 3/2 | 74 (54–84) | Stage I: 1<br>Stage II: |

| Disease source | Sex (female/male) | Median age (range), years | Disease stage |
|---|---|---|---|
| | | | Stage III: 1<br>Stage IV: 3<br>Stage X: |
| Renal (n = 5)<br>Indivumed | 1/4 | 70 (55–71) | Stage I: 2<br>Stage II:<br>Stage III:<br>Stage IV:<br>Stage X: 3 |
| Hepatic (n = 5)<br>Indivumed | 1/4 | 69 (56–81) | Stage I: 1<br>Stage II: 2<br>Stage III: 1<br>Stage IV: 1<br>Stage X: |
| Lung (n = 30)<br>Indivumed | 12/18 | 66 (52–82) | Stage I: 7<br>Stage II: 8<br>Stage III: 6<br>Stage IV: 9<br>Stage X: |
| Melanoma (n = 3)<br>Indivumed | 2/1 | 78 (71–79) | Stage I:<br>Stage II:<br>Stage III:<br>Stage IV:<br>Stage X: 3 |
| Ovarian (n = 5)<br>Indivumed | 5/0 | 68 (52–87) | Stage I: 1<br>Stage II:<br>Stage III: 3<br>Stage IV: 1<br>Stage X: |
| Pancreatic (n = 5)<br>Indivumed | 5/0 | 74 (59–89) | Stage I:<br>Stage II:<br>Stage III: 1<br>Stage IV: 4<br>Stage X: |
| Pharyngeal (n = 5)<br>Indivumed | 3/2 | 67 (58–78) | Stage I: 1<br>Stage II:<br>Stage III:<br>Stage IV: 1<br>Stage X: 3 |
| Prostate (n = 30)<br>Indivumed | 0/30 | 76 (55–86) | Stage I: 6<br>Stage II: 8<br>Stage III: 7<br>Stage IV: 9<br>Stage X: |
| Stomach (n = 5)<br>Indivumed | 1/4 | 71 (59–78) | Stage I:<br>Stage II:<br>Stage III:<br>Stage IV: 3<br>Stage X: 2 |
| Thyroid (n = 4)<br>Indivumed | 0/4 | 65 (58–79) | Stage I:<br>Stage II:<br>Stage III:<br>Stage IV: 3<br>Stage X: 1 |

**Supplementary Table S3**. Native and formaldehyde crosslinked results for cf-H3.1-nucleosome measurements.

**Table S3a. *Formaldehyde treatment of whole blood.*** Nucleosomes crosslinked in whole blood by sample collection in Streck cfDNA BCTs (crosslinked) or in EDTA BCTs (native) from 10 asymptomatic healthy volunteers, 49 diseased hospitalized patients with elevated levels of CRP (>5 mg/mL) and 25 patients diagnosed with a solid cancer. Change (%) calculated as Change (%) = 100 x [(Crosslinked result – Native result)/Native result].

| Whole blood sample from subject | Native (EDTA plasma) | Crosslinked (Streck plasma) | Change (%) |
|---|---|---|---|
| Asymptomatic healthy volunteers | 6.6 | 5.4 | -18% |
| | 13.2 | 9.4 | -29% |
| | 17.0 | 12.1 | -29% |
| | 11.9 | 9.4 | -21% |
| | 22.7 | 14.1 | -38% |
| | 13.6 | 11.7 | -14% |
| | 13.1 | 9.0 | -31% |
| | 9.2 | 6.5 | -29% |
| | 28.0 | 18.1 | -35% |
| | 11.5 | 8.0 | -30% |
| Diseased hospitalized patients with elevated levels of CRP (>5 mg/mL) | 207.5 | 87.7 | -58% |
| | 164.3 | 94.1 | -43% |
| | 32 | 16.8 | -48% |
| | 372.3 | 39.2 | -89% |
| | 384.9 | 290.4 | -25% |
| | 325.2 | 126.8 | -61% |
| | 343 | 185.7 | -46% |
| | 373 | 32.7 | -91% |
| | 410.6 | 129.3 | -69% |
| | 268.4 | 187.6 | -30% |
| | 173.3 | 53.1 | -69% |
| | 151.5 | 68.2 | -55% |
| | 114.3 | 55.4 | -52% |
| | 80.2 | 23.9 | -70% |
| | 91.3 | 61.8 | -32% |
| | 18.5 | 12.4 | -33% |
| | 43.3 | 11.2 | -74% |
| | 599.5 | 591.8 | -1% |
| | 110.8 | 61 | -45% |
| | 423.4 | 299.4 | -29% |
| | 99.8 | 44.2 | -56% |
| | 51.5 | 20.4 | -60% |
| | 480.1 | 293.2 | -39% |
| | 83.8 | 20.8 | -75% |
| | 121.9 | 15.2 | -88% |
| | 238.9 | 132 | -45% |
| | 77.5 | 36.5 | -53% |
| | 571.3 | 349.4 | -39% |
| | 121.8 | 77.1 | -37% |
| | 23.3 | 18.3 | -21% |

| **Whole blood sample from subject** | **Native (EDTA plasma)** | **Crosslinked (Streck plasma)** | **Change (%)** |
|---|---|---|---|
| | 68.3 | 36.1 | -47% |
| | 44.8 | 33.1 | -26% |
| | 242 | 94.1 | -61% |
| | 111.6 | 78.5 | -30% |
| | 27.9 | 13.7 | -51% |
| | 36.4 | 24.5 | -33% |
| | 174.4 | 92.5 | -47% |
| | 146 | 80.8 | -45% |
| | 112 | 48.5 | -57% |
| | 56.8 | 35.7 | -37% |
| | 101 | 62.7 | -38% |
| | 174.4 | 102.1 | -41% |
| | 234.3 | 138.8 | -41% |
| | 65.5 | 47 | -28% |
| | 65.7 | 46 | -30% |
| | 75.3 | 39.9 | -47% |
| | 252 | 204.3 | -19% |
| | 95.6 | 53.1 | -44% |
| | 62.8 | 24.8 | -61% |
| Ovarian cancer | 15.62 | 3.71 | -76% |
| CRC | 42.95 | 17.32 | -60% |
| CRC | 480.33 | 207.81 | -57% |
| CRC | 67.11 | 32.94 | -51% |
| Gastric cancer | 335.57 | 173.87 | -48% |
| CRC | 150.34 | 77.90 | -48% |
| Mesothelioma | 8.25 | 4.37 | -47% |
| CRC | 25.70 | 14.36 | -44% |
| CRC | 22.91 | 12.99 | -43% |
| Appendiceal cancer | 18.45 | 10.95 | -41% |
| CRC | 26.59 | 16.72 | -37% |
| CRC | 33.40 | 22.65 | -32% |
| CRC | 11.99 | 8.16 | -32% |
| Appendiceal cancer | 790.16 | 606.78 | -23% |
| CRC | 22.16 | 17.51 | -21% |
| CRC | 16.01 | 16.30 | +2% |
| CRC | 462.19 | 489.13 | +6% |
| CRC | 18.26 | 24.51 | +34% |
| CRC | 10.00 | 14.44 | +44% |
| Gastric cancer | 23.52 | 37.29 | +59% |
| CRC | 3.08 | 5.42 | +76% |
| CRC | 2.72 | 6.68 | +146% |
| Appendiceal cancer | 2.60 | 7.59 | +192% |
| CRC | 7.08 | 30.25 | +327% |
| CRC | 34.22 | 151.65 | +343% |

**Table S3b. *Formaldehyde treatment of plasma*.** Nucleosomes crosslinked in EDTA plasma samples from 40 patients diagnosed with CRC by treatment with 3.33% v/v Streck cfDNA BCT additive (crosslinked) or buffer (native). Change (%) calculated as Change (%) = 100 x [(Crosslinked result – Native result)/Native result].

| EDTA plasma sample from subject | Native (EDTA plasma treated with buffer) | Crosslinked (EDTA plasma treated with Streck additive) | Change (%) |
|---|---|---|---|
| CRC patients | 8.9 | 8.6 | -3% |
| | 31.2 | 28 | -10% |
| | 13.1 | 12.5 | -5% |
| | 20.2 | 19 | -6% |
| | 5 | 4.7 | -6% |
| | 19.5 | 19.1 | -2% |
| | 22.2 | 19.2 | -14% |
| | 16.1 | 15.1 | -6% |
| | 43.6 | 39.2 | -10% |
| | 24.4 | 21.9 | -10% |
| | 45 | 38.5 | -14% |
| | 36.6 | 30.5 | -17% |
| | 23.4 | 21.4 | -9% |
| | 32.9 | 30.7 | -7% |
| | 19.4 | 17.6 | -9% |
| | 15.4 | 14.3 | -7% |
| | 9.4 | 9.2 | -2% |
| | 27.8 | 25.5 | -8% |
| | 32.7 | 29.6 | -9% |
| | 54.1 | 48.6 | -10% |
| | 68.1 | 57.9 | -15% |
| | 150.8 | 131.2 | -13% |
| | 65.4 | 59.1 | -10% |
| | 130.8 | 116.3 | -11% |
| | 93.7 | 82.1 | -12% |
| | 215.9 | 165.1 | -24% |
| | 172.6 | 64.8 | -62% |
| | 99.5 | 87 | -13% |
| | 103.2 | 88.6 | -14% |
| | 69.6 | 58 | -17% |
| | 131.6 | 114.4 | -13% |
| | 80.8 | 68.7 | -15% |
| | 67.9 | 48.4 | -29% |
| | 104.8 | 90.8 | -13% |
| | 74.5 | 64 | -14% |
| | 88.8 | 59.5 | -33% |
| | 110.5 | 76.3 | -31% |
| | 79.8 | 64 | -20% |
| | 96.1 | 69.4 | -28% |
| | 86.1 | 73.8 | -14% |

**Supplementary Fig. S1.** Crosslinked cf-H3.1-nucleosome levels in asymptomatic healthy subjects analyzed at Volition by age (in decades). Data for 20 samples collected by Indivumed (red) and 70 patients collected by BioIVT (black). An additional 10 subjects from BioIVT were of unknown age.

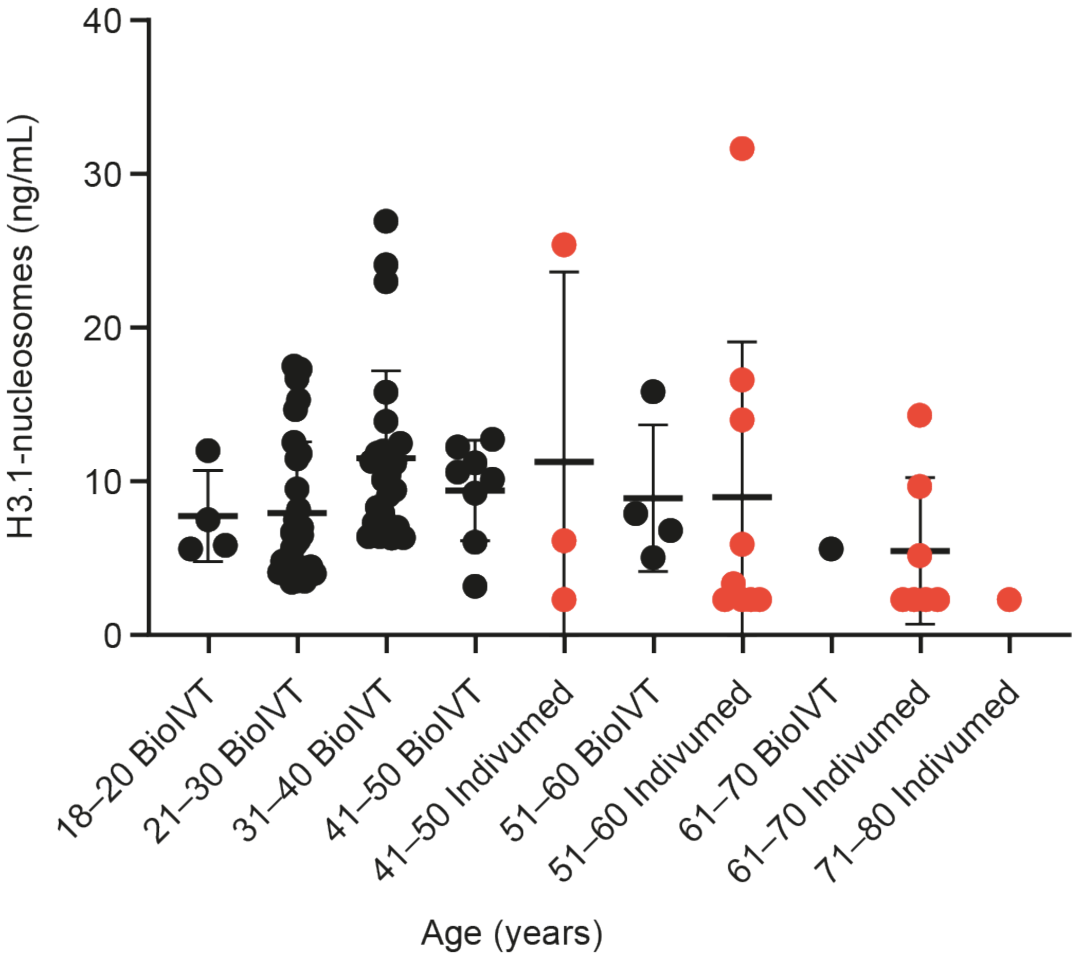

**Supplementary Fig. S2.** Frequency profile diagrams for four CRC Streck plasma samples.

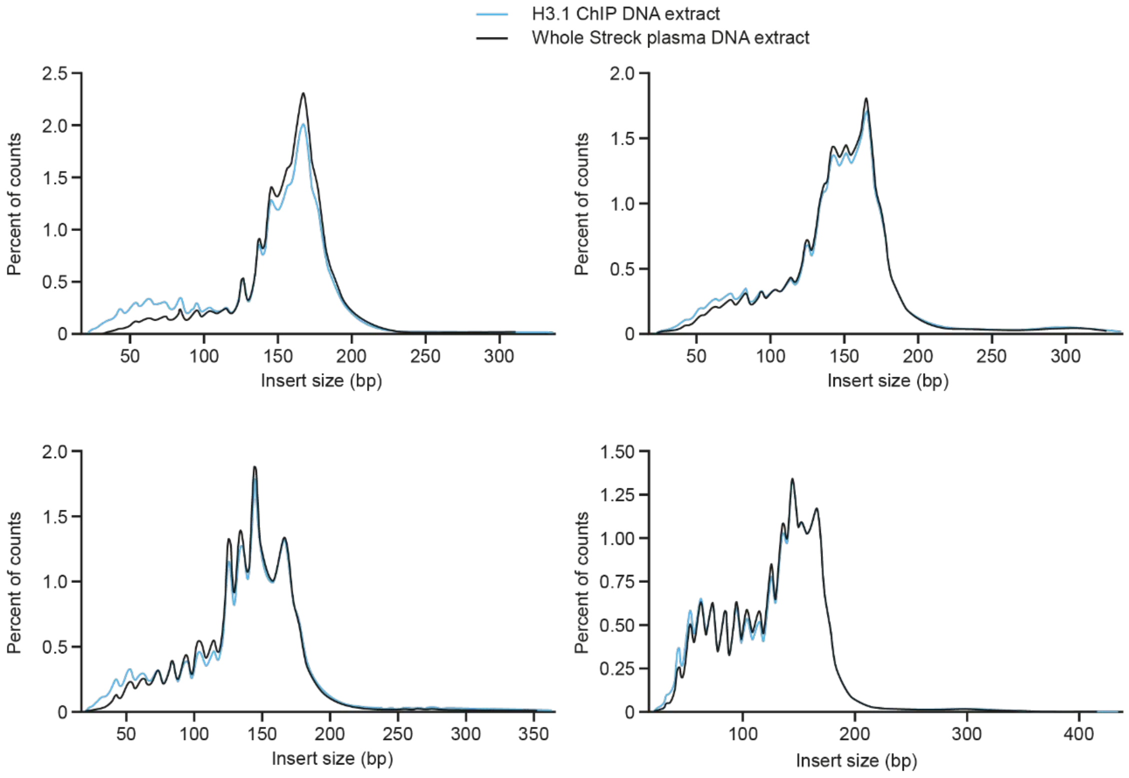
